# Sex-specific effects of aging on the humoral immune response to repeat vaccination with the high-dose seasonal influenza vaccine in older adults

**DOI:** 10.1101/2021.07.21.21260712

**Authors:** Janna R. Shapiro, Huifen Li, Rosemary Morgan, Yiyin Chen, Helen Kuo, Xiaoxuan Ning, Patrick Shea, Cunjin Wu, Katherine Merport, Rayna Saldanha, Suifeng Liu, Engle Abrams, Yan Chen, Denise C. Kelly, Eileen Sheridan-Malone, Lan Wang, Scott L. Zeger, Sabra L. Klein, Sean X. Leng

## Abstract

Older adults (≥65 years of age) bear a significant burden of severe disease and mortality associated with influenza, despite relatively high annual vaccination coverage and substantial pre-existing immunity to influenza. To test the hypothesis that host factors, including age and sex, play a role in determining the effect of repeat vaccination and levels of pre-existing humoral immunity to influenza, we evaluated pre- and post-vaccination strain-specific hemagglutination inhibition (HAI) titers in adults over 75 years of age who received a high-dose influenza vaccine in at least four out of six influenza seasons (NCT02200276). Neither age, sex, body mass index, frailty, nor repeat vaccination were significantly associated with post-vaccination HAI titer outcomes. Pre-vaccination titers, however, were significantly predictive of post-vaccination outcomes. Pre-vaccination titers to H1N1 remained constant with age, while those to H3N2 and influenza B decreased substantially with age in males but not in females. Our findings highlight the importance of pre-existing immunity in this highly vaccinated older adult population and suggest that older males are particularly vulnerable to reduced pre-existing humoral immunity to influenza from previous annual vaccination.

## Introduction

Seasonal influenza is an important public health burden in older adults (people ≥65 years of age), particularly the oldest and frail subset^1-3^. In the United States (U.S.), there are an estimated 4 million incident cases per year in older adults, accounting for 90% of deaths associated with influenza^4,5^. The U.S. Centers for Disease Control and Prevention (CDC) recommends annual influenza vaccination for prevention of influenza infection and complications in people 6 months and older^6^. The high-dose inactivated influenza vaccine (HD-IIV) is available to older adults and has demonstrated superior efficacy over standard-dose vaccines in this vulnerable age group^6,7^. Seasonal influenza vaccine coverage is relatively high in older adults, with >60% of older Americans vaccinated annually, compared to <40% of the 18-49 age group^8^.

Age-related immunosenescence, defined by a decline in cellular and humoral immune function combined with a chronic low-grade inflammatory phenotype (CLIP)^9-11^, is believed to be the primary reason for the reduced effectiveness of influenza vaccines observed in older adults^12-14^. Repeated annual vaccination may also have a negative effect on vaccine-induced humoral immune responses as well as vaccine effectiveness (VE). For example, a recent observational test-negative study using ten years of vaccination history found that in older adults, VE decreased with increasing numbers of previous vaccinations but that vaccination continued to offer some level of protection^15^. Another study over eight seasons in the general adult population found that VE to H3N2, but not influenza B, was reduced among individuals with frequent vaccination history compared to those without prior vaccination^16^. The impact of repeat vaccination on H3N2 was confirmed by a meta-analysis, which found heterogeneous effects of repeat vaccination overall; however, when negative effects were observed, they were most pronounced for H3N2^17^. In contrast, a recent systematic review and meta-analysis concluded that the available evidence does not support a reduction in VE with consecutive repeat vaccination, but that certainty in the evidence was low^18^. Case control studies in both Australia and Spain found beneficial effects of repeated annual vaccination on VE in older adults^19,20^, and an observational population-based study in Sweden found no differences in VE between those who had been vaccinated in the current season only and those who had been vaccinated in both the current and previous seasons^21^. Based on the conflicting evidence, multi-season clinical studies to address the effects of aging and repeated vaccination have been recommended^17^.

Mechanistically, pre-existing immunity generated to various influenza virus exposures over time can have an important impact on the outcome of vaccination. According to immune imprinting theory, the memory response established by an individual’s first influenza exposure has a lifelong effect on subsequent immune responses to infection or vaccination^22^. Broad pre-existing immunity is thought to have negative consequences, as pre-existing antibodies can suppress the response to novel influenza virus strains by reducing the amount of available antigen or epitope masking^23,24^. A theoretical benefit of HD-IIV is that pre-existing antibodies cannot sequester the increased amount of antigen delivered, and, thus, more antigen is available to activate memory B cells and elicit a protective response^25-27^. To our knowledge, however, the impact of pre-existing immunity in the context of the HD-IIV has not been adequately characterized.

In addition to age, other host factors including sex, frailty, and body mass index (BMI) can impact vaccine responses in older adults. Females have been found to mount greater antibody responses to HD-IIV than males^28^. The relationship between frailty and influenza vaccine responses is debated in the literature, with one study reporting frailty having a negative effect^29^, others reporting no effect^30-34^, and others still reporting a positive effect^35^. Finally, in older adults, obesity is significantly associated with decreased hemagglutination inhibition (HAI) titers and percentage of switched memory B cells^36^. We hypothesize that the variation across studies in estimates of the effects of pre-existing immunity and repeat annual vaccination is partly caused by failure to adequately account for heterogeneity and interactions among host factors that likely differ across studies. To address this knowledge gap, we used a longitudinal cohort of older adults over 75 years of age who had received high-dose, trivalent inactivated influenza vaccine (HD-IIV3) in at least four out of six influenza seasons to estimate the impact of repeat vaccination on the antibody response to HD-IIV3 and its dependence on the intersection of age, sex, frailty, BMI, pre-existing immunity.

## Results

### Study Participants and annual influenza immunization with HD-IIV3

Over the six influenza seasons from 2014-2015 to 2019-2020, 90 individuals participated in at least four study seasons and 433 doses of HD-IIV3 were. The strains included in each vaccine and the study protocol are described in **Figure 1. Table 1** shows demographic and clinical characteristics of the study participants. There was a slight predominance of females (55.6%), and yearly study enrollment increased over time. Baseline characteristics, measured during the first year of participation, were similar between males and females. The median age at study enrollment was 80, and >50% of participants were classified as pre-frail as per the Frailty Phenotype^37^. Trends in the change in frailty since baseline, calculated as the difference between the first and last years of participation, differed by sex. A greater proportion of males improved in frailty status, whereas more females either did not change or progressed in frailty status. A greater proportion of males also experienced changes in BMI over the course of the study, while BMI did not change for females.

**Figure 1.**
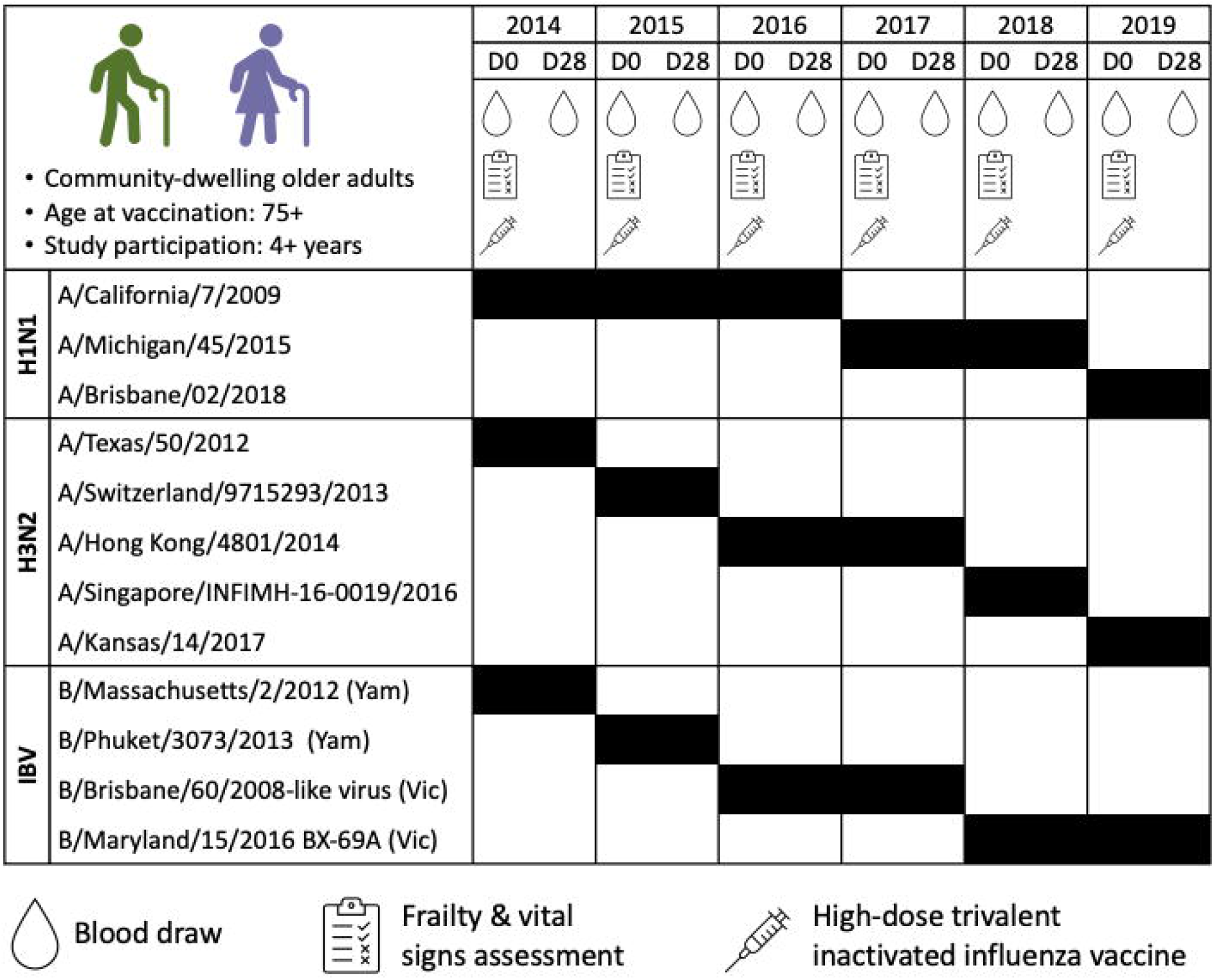
Study design. Study procedures and the three strains included in each seasonal HD-IIV3 are shown. Serum from blood draws was used to evaluate pre- and post-vaccination strain-specific hemagglutination antibody inhibition (HAI) titers, and frailty was assessed using the Frailty Phenotype.

**Table 1.**
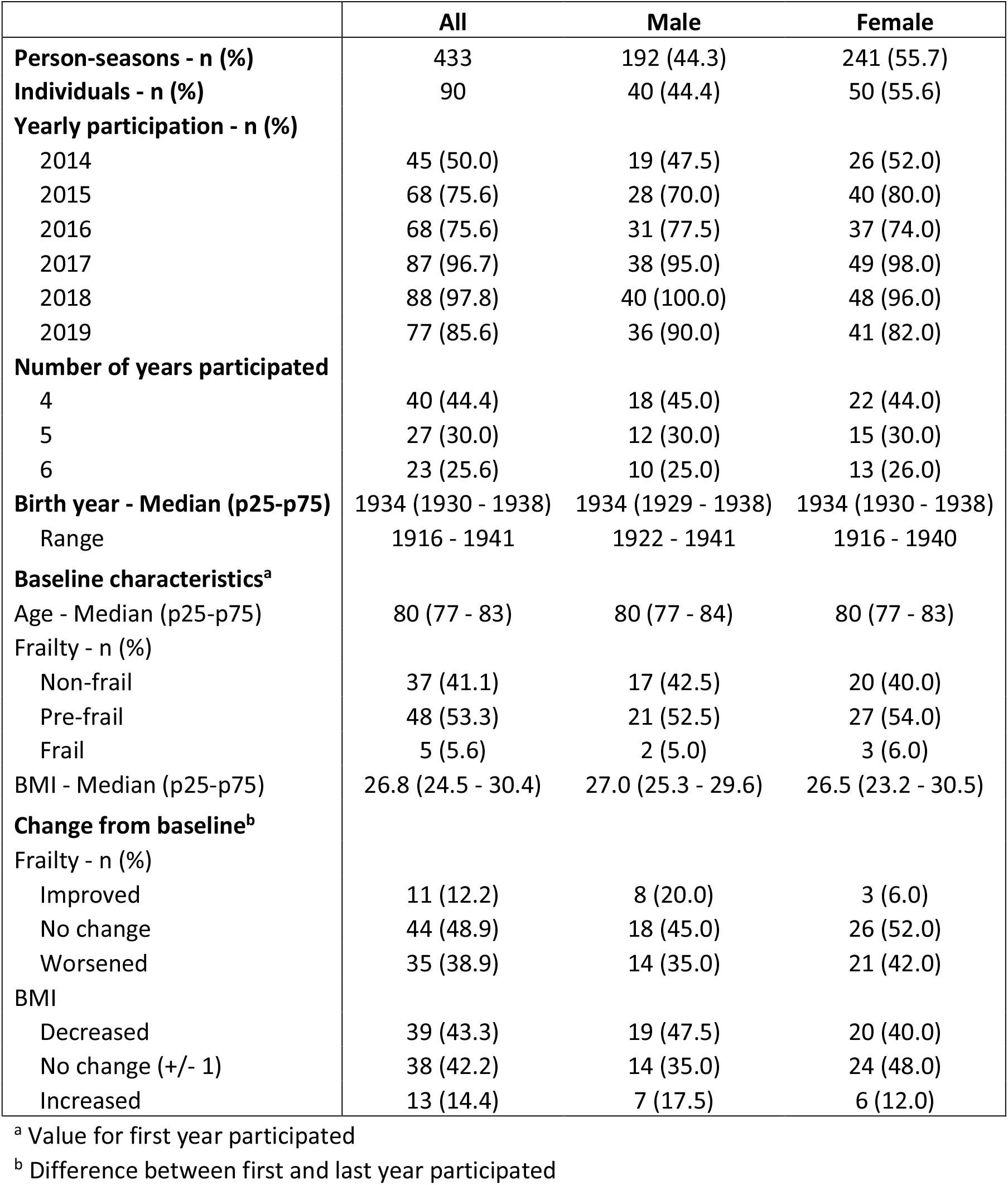
Summary of study population characteristics.

### Pre- and post-vaccination strain-specific HAI titers are high among repeatedly vaccinated older adults

Pre- and post-vaccination strain-specific HAI titer outcomes to the three HD-IIV3 vaccine antigens are summarized and disaggregated by sex in **Table 2**. As expected in this highly vaccinated elderly population, titers and seroprotection rates (defined as an HAI titer ≥40^38-40^) were high. This was particularly true for influenza B, where 98% of the participants were seroprotected prior to immunization. Post-vaccination, >94% of participants achieved seroprotection for H1N1 and H3N2, while 100% of participants were seroprotected against influenza B. Because of the lack of variability in post-vaccination seroprotection, this outcome was omitted from further analysis. Fold-rise in titers and rates of seroconversion, as defined by ≥4-fold titer increase^41^, were relatively low but were highest for H3N2. There were no sex differences in any post-vaccination outcomes. Together, these data indicate that pre-existing and post-vaccination strain-specific HAI titers remained high among older adults who were repeatedly vaccinated with HD-IIV3.

**Table 2.**
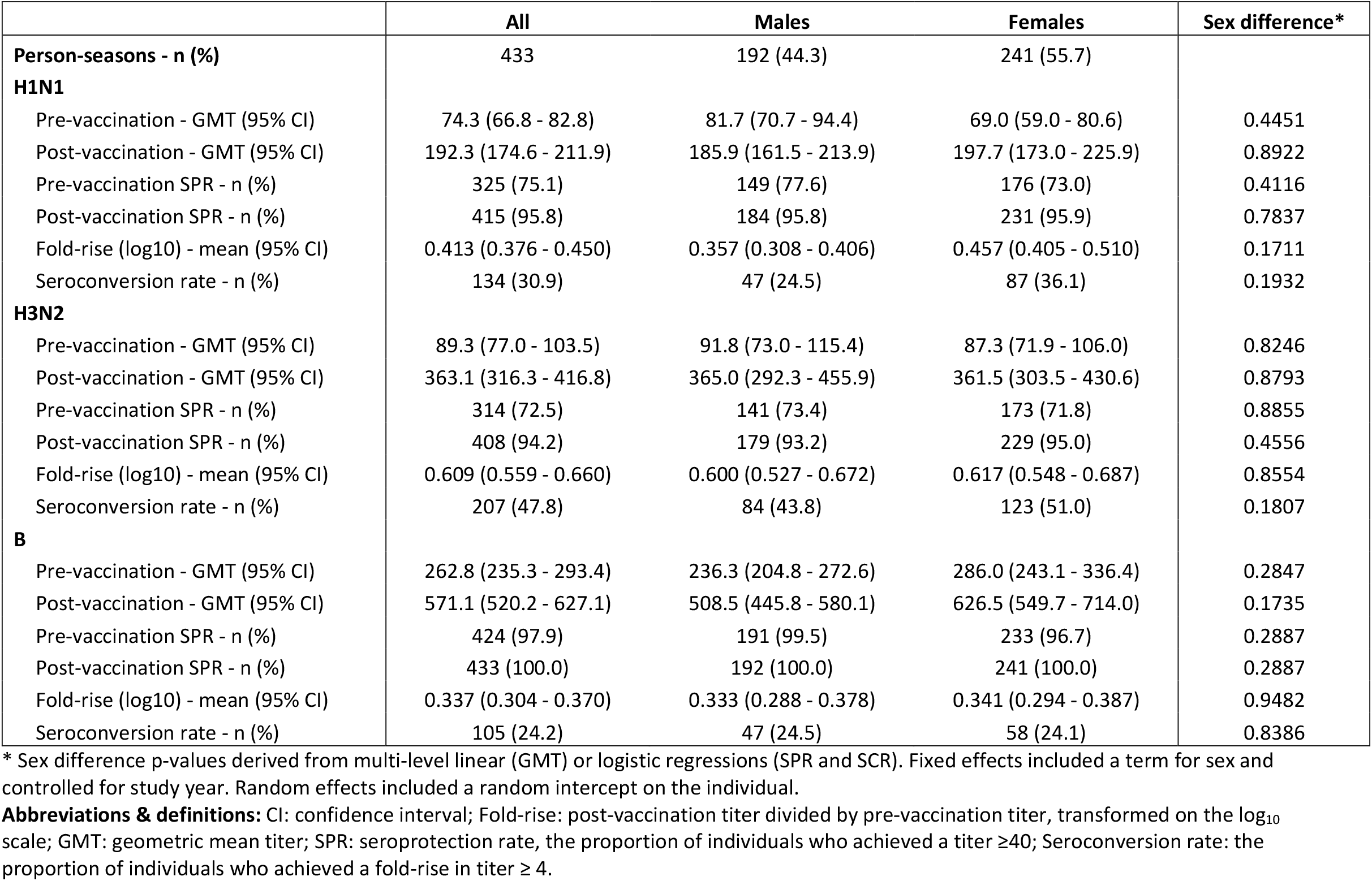
Pre- and post-vaccination hemagglutination antibody inhibition (HAI) titer outcomes.

|  | All | Males | Females | Sex difference* |
| --- | --- | --- | --- | --- |
| **Person-seasons - n (%)** | 433 | 192 (44.3) | 241 (55.7) |  |
| **H1N1** |  |  |  |  |
| Pre-vaccination - GMT (95% CI) | 74.3 (66.8 - 82.8) | 81.7 (70.7 - 94.4) | 69.0 (59.0 - 80.6) | 0.4451 |
| Post-vaccination - GMT (95% CI) | 192.3 (174.6 - 211.9) | 185.9 (161.5 - 213.9) | 197.7 (173.0 - 225.9) | 0.8922 |
| Pre-vaccination SPR - n (%) | 325 (75.1) | 149 (77.6) | 176 (73.0) | 0.4116 |
| Post-vaccination SPR - n (%) | 415 (95.8) | 184 (95.8) | 231 (95.9) | 0.7837 |
| Fold-rise (log10) - mean (95% CI) | 0.413 (0.376 - 0.450) | 0.357 (0.308 - 0.406) | 0.457 (0.405 - 0.510) | 0.1711 |
| Seroconversion rate - n (%) | 134 (30.9) | 47 (24.5) | 87 (36.1) | 0.1932 |
| **H3N2** |  |  |  |  |
| Pre-vaccination - GMT (95% CI) | 89.3 (77.0 - 103.5) | 91.8 (73.0 - 115.4) | 87.3 (71.9 - 106.0) | 0.8246 |
| Post-vaccination - GMT (95% CI) | 363.1 (316.3 - 416.8) | 365.0 (292.3 - 455.9) | 361.5 (303.5 - 430.6) | 0.8793 |
| Pre-vaccination SPR - n (%) | 314 (72.5) | 141 (73.4) | 173 (71.8) | 0.8855 |
| Post-vaccination SPR - n (%) | 408 (94.2) | 179 (93.2) | 229 (95.0) | 0.4556 |
| Fold-rise (log10) - mean (95% CI) | 0.609 (0.559 - 0.660) | 0.600 (0.527 - 0.672) | 0.617 (0.548 - 0.687) | 0.8554 |
| Seroconversion rate - n (%) | 207 (47.8) | 84 (43.8) | 123 (51.0) | 0.1807 |
| **B** |  |  |  |  |
| Pre-vaccination - GMT (95% CI) | 262.8 (235.3 - 293.4) | 236.3 (204.8 - 272.6) | 286.0 (243.1 - 336.4) | 0.2847 |
| Post-vaccination - GMT (95% CI) | 571.1 (520.2 - 627.1) | 508.5 (445.8 - 580.1) | 626.5 (549.7 - 714.0) | 0.1735 |
| Pre-vaccination SPR - n (%) | 424 (97.9) | 191 (99.5) | 233 (96.7) | 0.2887 |
| Post-vaccination SPR - n (%) | 433 (100.0) | 192 (100.0) | 241 (100.0) | 0.2887 |
| Fold-rise (log10) - mean (95% CI) | 0.337 (0.304 - 0.370) | 0.333 (0.288 - 0.378) | 0.341 (0.294 - 0.387) | 0.9482 |
| Seroconversion rate - n (%) | 105 (24.2) | 47 (24.5) | 58 (24.1) | 0.8386 |

\* Sex difference p-values derived from multi-level linear (GMT) or logistic regressions (SPR and SCR). Fixed effects included a term for sex and controlled for study year. Random effects included a random intercept on the individual.
**Abbreviations & definitions:** CI: confidence interval; Fold-rise: post-vaccination titer divided by pre-vaccination titer, transformed on the log<sub>10</sub> scale; GMT: geometric mean titer; SPR: seroprotection rate, the proportion of individuals who achieved a titer ≥40; Seroconversion rate: the proportion of individuals who achieved a fold-rise in titer ≥ 4.

### Age, sex, BMI, frailty status and repeat vaccination are not associated with post-vaccination outcomes

We then assessed the relationships between pre-defined host factors (i.e., age, sex, BMI, and frailty status) and post-vaccination strain-specific HAI titer outcomes. When controlling for pre-vaccination titers and study year, neither age, frailty, nor BMI were individually statistically-significantly associated with post-vaccine titers (**Figure 2A-C**), the fold-rise in titers (**Figure 2D-F**), or the odds of seroconversion (**Figure 2G-I**) for either H1N1, H3N2 or influenza B. Inclusion of interaction terms in the models allowed for analysis of sex-specific contributions of age, frailty, and BMI as well as sex differences in the effects of these host factors. None of the host factors had a statistically significant association with post-vaccination outcomes for either males or females, and no statistically significant sex differences in post-vaccination outcomes were observed.

**Figure 2.**
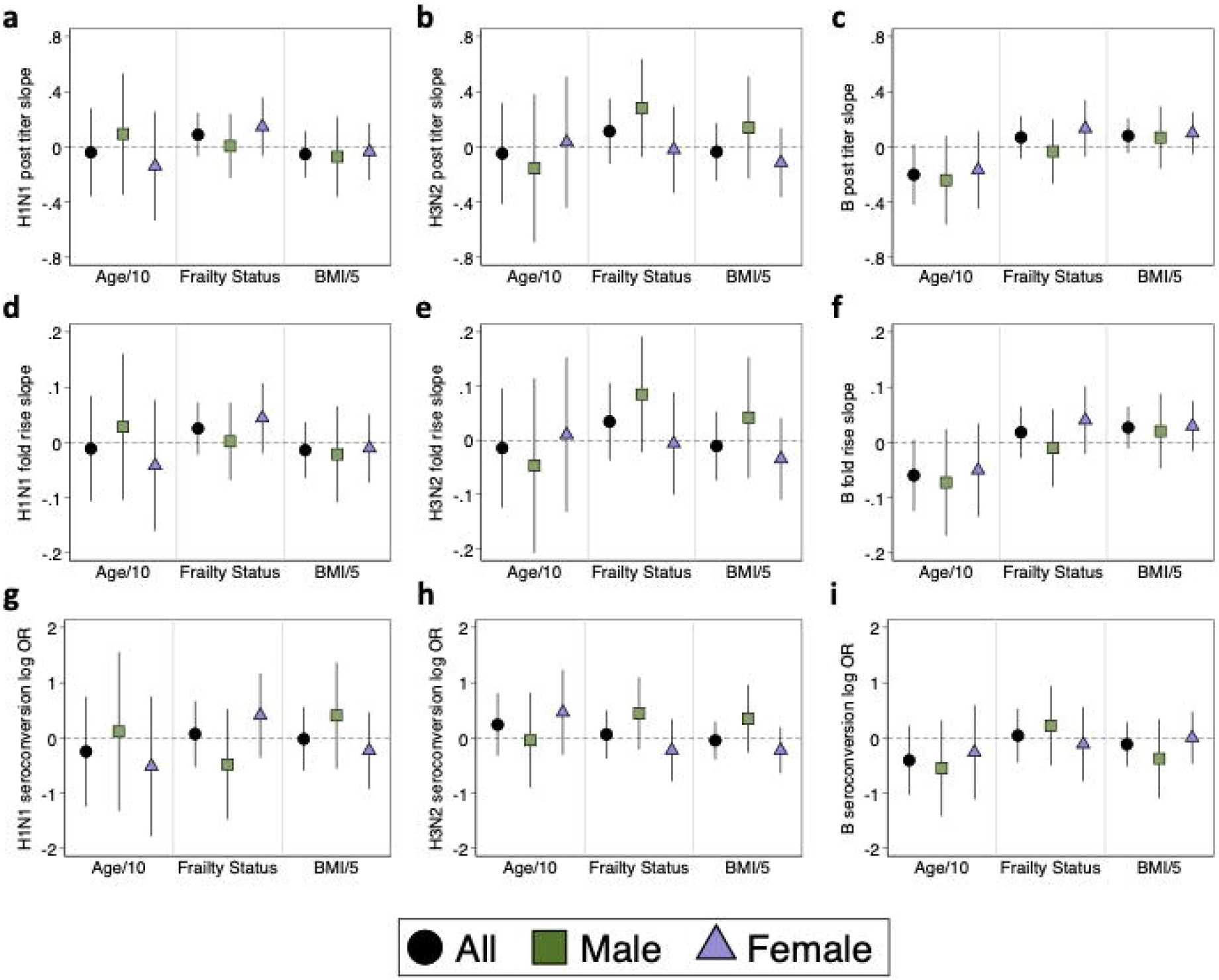
Relationship of age, frailty status, and BMI with post-vaccination hemagglutination antibody inhibition (HAI) titers outcomes. The relationship of age (in decades, Age/10), frailty status, and BMI (five-unit intervals, BMI/5) with log_2_-transformed post-vaccination titers are shown as slopes for HAI titers against H1N1 (**a**), H3N2 (**b**) and influenza B (**c**). The relationships of these host factors with log_10_-transformed fold-rise in titers (post-titer/pre-titer) are shown as slopes for H1N1 (**d**), H3N2 (**e**) and influenza B (**f**). Their relationships with log odds of seroconversion are shown for H1N1 (**g**), H3N2 (**h**) and influenza B (**i**). Estimates and 95% confidence intervals were derived from multi-level mixed effects models with random intercepts on the individual participant. Models controlled for study year and pre-vaccination HAI titers, and either controlled for sex (whole population estimates) or used interaction terms between sex and the host factor of interest to derive sex-specific estimates.

The relationship between the number of years of study participation and post-vaccination HAI titers outcomes was investigated to address the concern of potential negative effects of repeated annual vaccination on the humoral immune response to future vaccines. Vaccination status prior to study enrollment could not be verified, but leveraging the longitudinal design of our study, we sought to quantify whether post-vaccination outcomes declined with additional annual vaccination, and whether this effect differed by sex. There was generally a negative trend between the number of years participated and post-vaccination titers (**Figure 3A-D**), fold rise in titers (**Figure 3E-H**), and odds of seroconversion (**Figure 3I-L**); these decreases, however, were not statistically significant.

**Figure 3.**
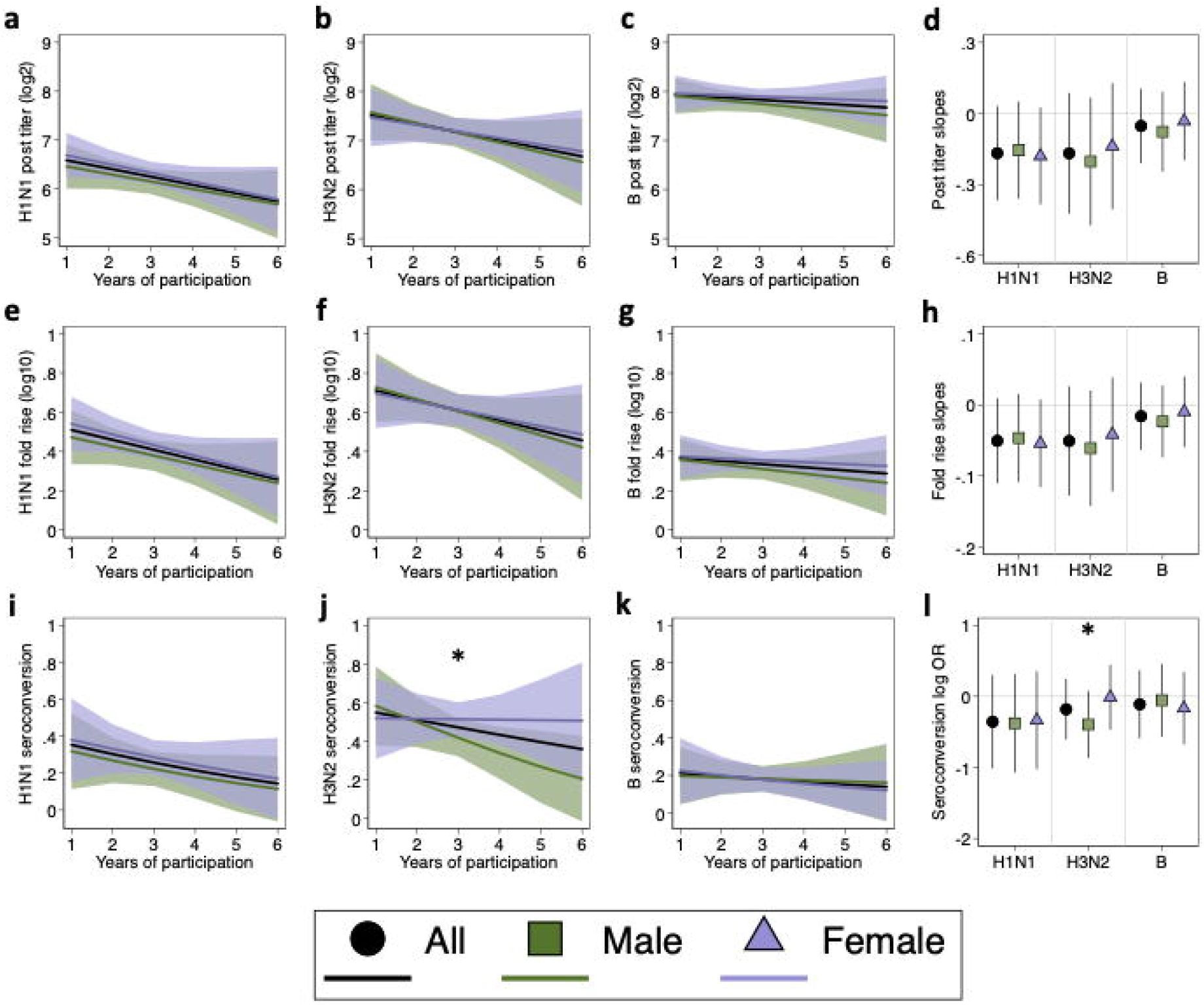
Relationship of repeat annual vaccination with post-vaccination hemagglutination antibody inhibition (HAI) titer outcomes. The relationship between increasing years of vaccination and log_2_-transformed post-vaccination HAI titers are shown for each vaccine antigen (**a-c**), with slopes summarized (**d**). Similarly, the relationship between increasing years of vaccination and the log_10_-transformed fold-rise in titers are shown for H1N1, H3N2, and influenza B (**e-g**), with the slopes summarized (**h**). The relationship between increasing years of vaccination and probability of seroconversion are shown for antibody responses against H1N1, H3N2, and influenza B, respectively (**i-k**), with the logs odds ratios summarized (**I**). All models controlled for study year, pre-vaccination titers, and age at vaccination. Whole-population models controlled for sex, while sex-specific estimates were derived from models including an interaction term between sex and years of participation and are shown with 95% confidence intervals. Asterisks indicate significant sex differences.

Two interesting trends emerged from this analysis. First, the effect sizes were smaller (i.e., closer to the null) for influenza B than for either H1N1 or H3N2, suggesting that the impact of repeat vaccination may vary by influenza subtype. Secondly, there was a significant sex difference in the effect of increasing years of vaccination on the odds of seroconverting to H3N2 (difference in log odds = 0.38; p = 0.023). While there was no effect in females (OR = 0.988; p = 0.959), the odds of seroconverting to H3N2 decreased by 33% for each additional year of study participation for males (OR = 0.673; p = 0.100) (**Figure 3K-L**). Therefore, in addition to varying by virus subtype, the mechanisms through which repeated vaccination affects antibody responses may be sex-specific.

### Pre-vaccination HAI titers strongly predict post-vaccination outcomes

Overall, host factors and increasing years of vaccination did not significantly predict any of the post-vaccination antibody titer parameters. Pre-vaccination titers, however, were strong predictors of post-vaccination titers for H1N1 (slope = 0.49; 95% CI: 0.41; 0.58) (**Figure 4A**), H3N2 (slope = 0.59; 95% CI: 0.52; 0.66) (**Figure 4B**), and influenza B (slope = 0.57; 95% CI: 0.50; 0.63) (**Figure 4C**) (p<0.0001 for testing the null hypotheses that the slope is zero for each antigen). Conversely, there was a strong negative association between pre-vaccination titers and the fold-rise for H1N1 (slope = -0.15; 95% CI: 0.18; 0.13) (**Figure 4D**), H3N2 (slope = -0.12; 95% CI: -0.15; -0.10) (**Figure 4E**), and influenza B (slope = -0.13; 95% CI: -0.15; -0.11) (**Figure 4F**) (p<0.0001 for testing the null hypotheses that the slope to zero for all antigens), such that greater pre-vaccination titers were associated with a smaller fold rise. Finally, there was a similar negative association between pre-vaccination titers and the odds of seroconverting for H1N1 (OR = 0.31; 95% CI: 0.21; 0.46) (**Figure 4G**), H3N2 (OR = 0.60; 95% CI: 0.50; 0.71) (**Figure 4H**), and influenza B (OR = 0.41; 95% CI: 0.31; 0.55) (**Figure 4I**) (p<0.0001 for testing the null hypothesis that the odds ratio is equal to one for all antigens).

**Figure 4.**
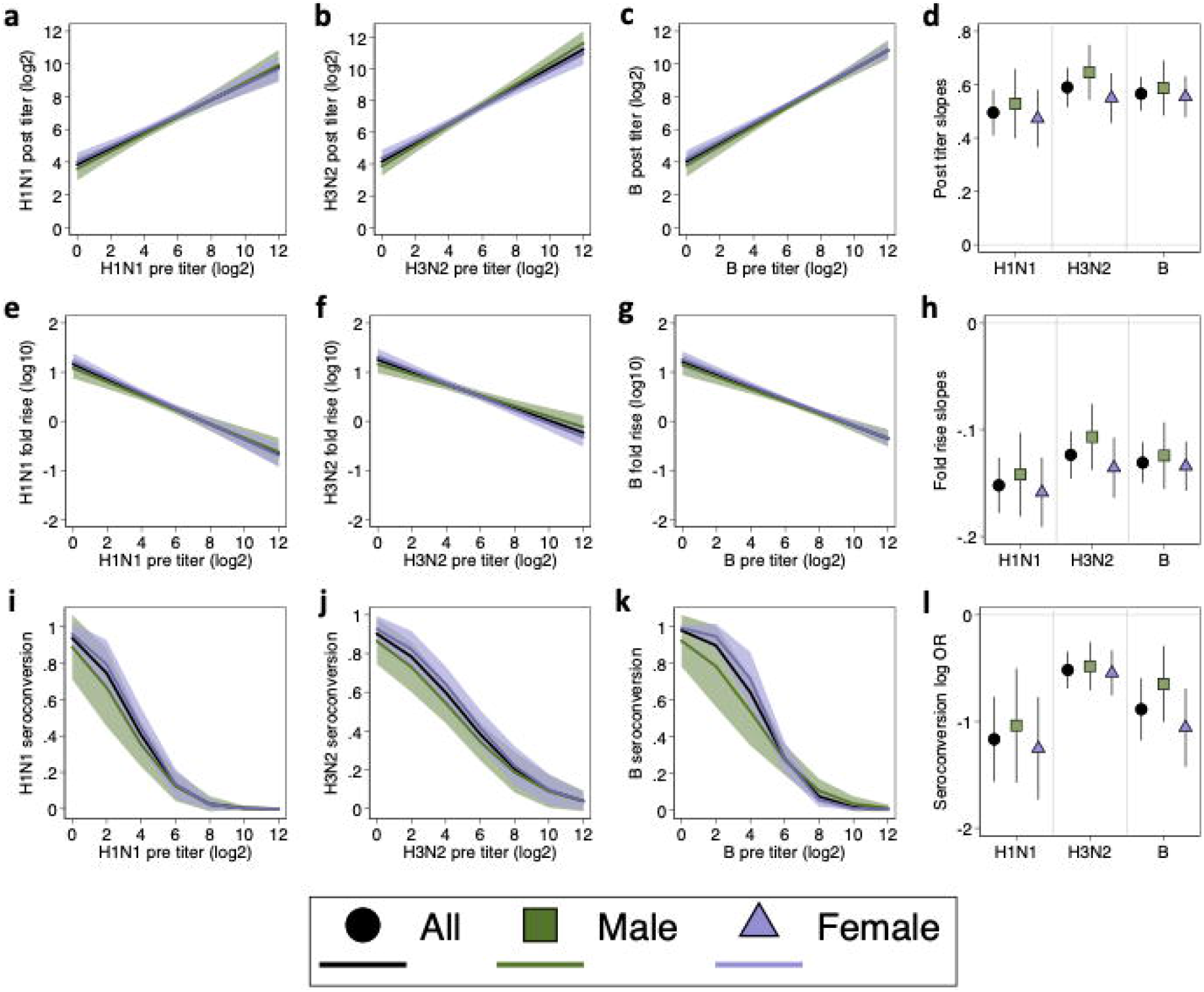
Relationship between pre-vaccination hemagglutination antibody inhibition (HAI) titers and post-vaccination HAI titer outcomes. Relationships between log_2_-transformed pre-vaccination HAI titers and log_2_-transformed post-vaccination titers are shown for responses to H1N1, H3N2, and influenza B, respectively (**a-c**), with slopes summarized (**d**). The relationships between pre-vaccination HAI titers and the log_10_-transformed fold-rise in titers are shown for each vaccine antigen (**e-g**), with the slopes summarized (**h**). The relationships between pre-vaccination HAI titers and the probability of seroconversion are shown for H1N1, H3N2, and influenza B, respectively (**i-k**), with the log odds ratios summarized (**l**). All models controlled for study year. Overall estimates controlled for sex, while sex-specific estimates were derived from models that included an interaction term between sex and pre-vaccination titers and are shown with 95% confidence intervals.

The strength of these associations suggests that post-vaccination outcomes are primarily determined by pre-existing humoral immunity. Thus, in highly vaccinated populations, such as the older adult participants in this study, pre-vaccination titers are not just confounders to be controlled for in the analysis of post-vaccination humoral immunity but are an outcome of public health importance that illustrate durability of immunity to influenza from one season to the next. Given the importance of pre-vaccination titers, we focused subsequent analyses on exploring the relationships between host factors and pre-vaccination HAI titers in the context of repeat annual vaccination.

### Sex modifies the relationship between age and pre-vaccination HAI titers

Next, we assessed the relationships between age, frailty, BMI, and pre-vaccination titers (**Figure 5A-C**). Neither frailty nor BMI were statistically significantly associated with pre-vaccination HAI titers for all participants or in sex-disaggregated subgroup analysis. Further, there were no statistically significant sex differences in the effects of frailty or BMI on pre-vaccination HAI titers against either H1N1, H3N2, or influenza B. A statistically significant sex by age interaction, however, was observed for H3N2 (**Figure 5B**) and for influenza B (**Figure 5C**), in which HAI titers declined with age among male but not female participants.

**Figure 5.**
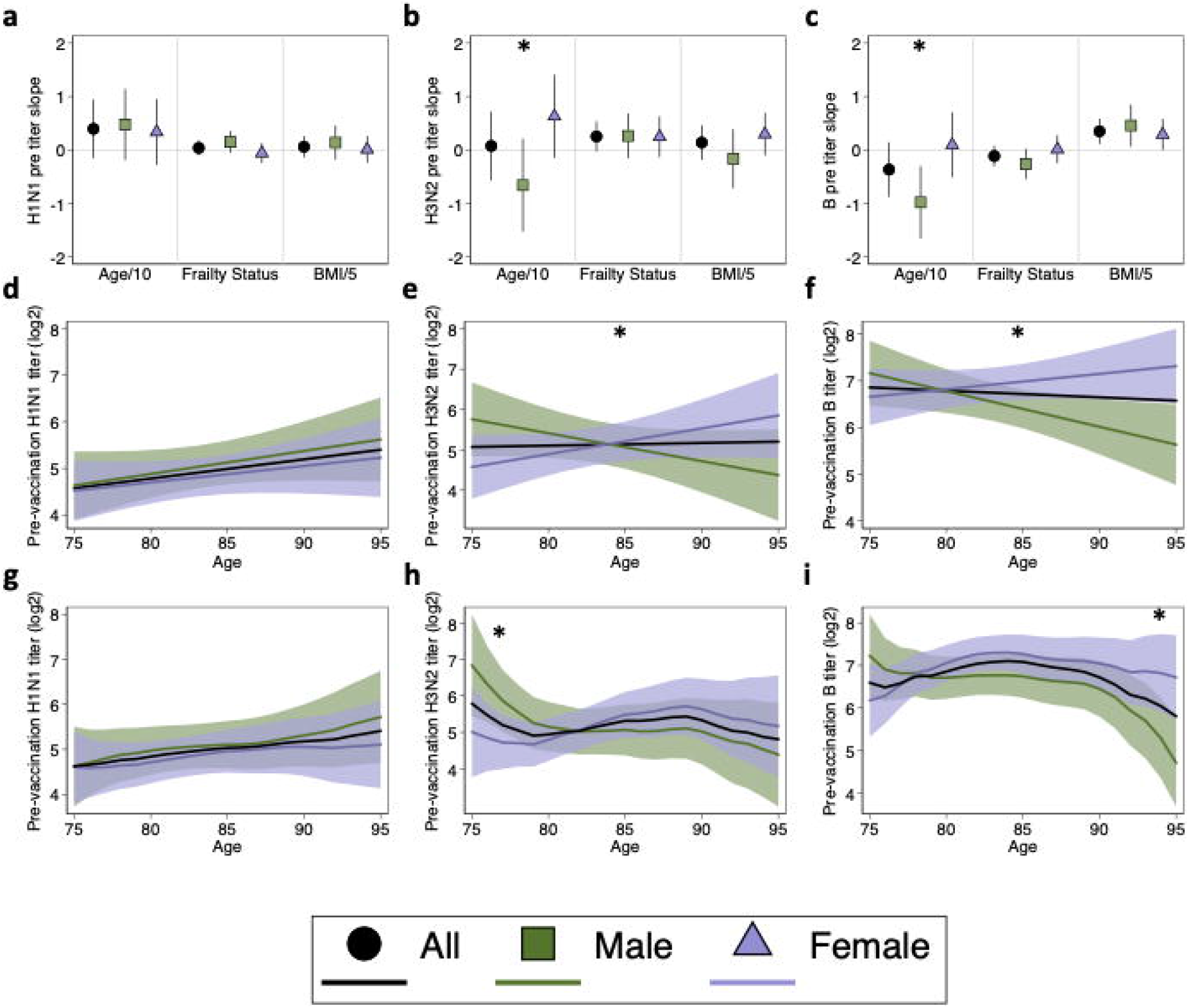
Relationship of age, frailty status, and BMI to pre-vaccination hemagglutination antibody inhibition (HAI) titers. Estimates for the relationship of age in decades (Age/10), frailty status, and BMI (five-unit intervals, BMI/5) to pre-vaccination HAI titers were derived from multilevel mixed-effects models controlling for study year for H1N1 (**a**), H3N2 (**b**), and influenza B (**c**). Expanded age models controlling for frailty and BMI are shown for responses to H1N1 (**d**), H3N2 (**e**), and influenza B (**f**). Expanded models for responses to H1N1 (**g**), H3N2 (**h**), and influenza B (**I**) were then amended to include cubic B-splines for age with knots at 5-year intervals. Models for the whole study population adjusted for sex, while sex-specific estimates included an interaction term allowing effects to differ by sex and are shown with 95% confidence intervals. Asterisks indicate significant sex differences.

To further interrogate the sex-specific effects of aging, expanded models controlling for frailty and BMI, in addition to study year, were constructed. Coefficients from the base (i.e., controlling for study year only) and expanded models are shown in **Table 3**, and results from expanded models are plotted in **Figure 5D-F**. For H1N1, HAI titers tended to increase with age for both males and females, but the increase was not statistically significant (males: 0.49 units per decade, p = 0.152; females: 0.35 units per decade, p = 0.267), nor was the sex difference in slope (p = 0.676) (**Figure 5D**). For H3N2, while titers again tended to increase with age in females (0.62 units per decade, p = 0.121), there was a decreasing trend with age in males (−0.75 units per decade, p =0.097), leading to a significant sex difference in age slopes (sex by age interaction = 0.137; p = 0.010) (**Figure 5E**). For influenza B, titers again tended to increase with age in females (0.33 units per decade, p = 0.275) but decreased by 0.78 units per decade in males (p = 0.023) (**Figure 5F**). Like H3N2, the sex difference in the effect of age was significant for influenza B (p = 0.005).

**Table 3.** Sex-specific effects of age on pre-vaccination hemagglutination antibody inhibition (HAI) titers.

|  | Base Models<sup>a</sup> |  |  |  |  |  | Expanded Models<sup>b</sup> |  |  |  |  |  |
| --- | --- | --- | --- | --- | --- | --- | --- | --- | --- | --- | --- | --- |
|  | Male age effects |  | Female age effects |  | Sex difference<sup>c</sup> |  | Male age effects |  | Female age effects |  | Sex difference<sup>c</sup> |  |
|  | Change | p-value | Change | p-value | Difference | p-value | Change | p-value | Change | p-value | Difference | p-value |
| **H1N1** |  |  |  |  |  |  |  |  |  |  |  |  |
| Linear | 0.047 | 0.164 | 0.034 | 0.286 | -0.014 | 0.674 | 0.049 | 0.152 | 0.035 | 0.267 | -0.014 | 0.676 |
| Non-linear |  |  |  |  |  |  |  |  |  |  |  |  |
| 75-80 | 0.379 | 0.300 | 0.146 | 0.650 | -0.023 | 0.963 | 0.383 | 0.301 | 0.134 | 0.685 | -0.010 | 0.984 |
| 80-85 | 0.133 | 0.564 | 0.233 | 0.251 | -0.256 | 0.465 | 0.131 | 0.574 | 0.243 | 0.238 | -0.259 | 0.462 |
| 85-90 | 0.205 | 0.397 | 0.109 | 0.589 | -0.156 | 0.657 | 0.220 | 0.377 | 0.113 | 0.593 | -0.147 | 0.678 |
| 90-95 | 0.421 | 0.188 | 0.066 | 0.830 | -0.253 | 0.547 | 0.430 | 0.186 | 0.071 | 0.817 | -0.254 | 0.553 |
| 95 |  |  |  |  | -0.609 | 0.31 |  |  |  |  | -0.612 | 0.313 |
| **H3N2** |  |  |  |  |  |  |  |  |  |  |  |  |
| Linear | -0.067 | 0.133 | 0.065 | 0.103 | **0.131** | **0.014** | -0.075 | 0.097 | 0.062 | 0.121 | **0.137** | **0.010** |
| Non-linear |  |  |  |  |  |  |  |  |  |  |  |  |
| 75-80 | **-1.725** | **0.015** | -0.156 | 0.801 | **-1.898** | **0.029** | **-1.776** | **0.012** | -0.297 | 0.637 | **-1.833** | **0.036** |
| 80-85 | -0.139 | 0.711 | **0.653** | **0.039** | -0.328 | 0.469 | -0.102 | 0.786 | **0.670** | **0.037** | -0.355 | 0.435 |
| 85-90 | 0.023 | 0.953 | 0.114 | 0.716 | 0.463 | 0.326 | -0.048 | 0.904 | 0.149 | 0.651 | 0.417 | 0.379 |
| 90-95 | -0.496 | 0.39 | -0.415 | 0.430 | 0.553 | 0.351 | -0.591 | 0.307 | -0.421 | 0.431 | 0.614 | 0.308 |
| 95 |  |  |  |  | 0.634 | 0.503 |  |  |  |  | 0.784 | 0.412 |
| **B** |  |  |  |  |  |  |  |  |  |  |  |  |
| Linear | **-0.098** | **0.005** | 0.009 | 0.762 | **0.108** | **0.007** | **-0.078** | **0.023** | 0.033 | 0.275 | **0.111** | **0.005** |
| Non-linear |  |  |  |  |  |  |  |  |  |  |  |  |
| 75-80 | -0.315 | 0.516 | **0.965** | **0.023** | -1.051 | 0.087 | -0.189 | 0.693 | **1.203** | **0.004** | -1.048 | 0.080 |
| 80-85 | -0.106 | 0.695 | 0.161 | 0.486 | 0.229 | 0.511 | 0.038 | 0.886 | 0.219 | 0.330 | 0.344 | 0.298 |
| 85-90 | -0.291 | 0.306 | **-0.534** | **0.019** | 0.496 | 0.165 | -0.181 | 0.515 | -0.111 | 0.633 | 0.525 | 0.123 |
| 90-95 | **-1.799** | **0.000** | -0.434 | 0.248 | 0.253 | 0.572 | **-1.649** | **0.000** | -0.233 | 0.526 | 0.595 | 0.168 |
| 95 |  |  |  |  | **1.618** | **0.020** |  |  |  |  | **2.011** | **0.003** |

<sup>a</sup> Base models controlled for study year.
<sup>b</sup> Expanded models controlled for study year, frailty, and BMI.
<sup>c</sup> For linear models, the sex-difference is the age-sex interaction term. For non-linear models, the sex difference is at the beginning of each five-year interval.
Non-linear models include cubic B-splines with knots at 5-year age intervals from 75-95 years.
Statistically significant values are bolded.

Both the base and expanded models were then amended to include cubic splines to obtain more granular estimates of the effects of age on HAI titers for males and females. Coefficients for both base and expanded non-linear models are shown in **Table 3**, and results from expanded models are plotted in **Figure 5G-I**. The non-linear model for H1N1 did not differ from the linear model, and no significant effects of age within each sex or difference between the sexes were observed (**Figure 5G**). Although the trends in the linear models were similar for H3N2 and influenza B, using age as a non-linear predictor revealed that different age categories were driving the overall effects. For H3N2, the increase in pre-vaccination HAI titers with age in females was driven by individuals in the 80-85 age category (increase of 0.67 units; p = 0.037), and the decrease in males was driven primarily by people in the 75-80 age category (decrease of 1.78 units; p = 0.012) (**Figure 5.H**). Thus, the sex difference was greatest at the younger end of the cohort (p = 0.036 at 75 years of age). Conversely, increasing titers to influenza B with age in females were driven by individuals in the 75-80 age category (increase of 1.2 units; p = 0.004), whereas for males there was a sharp decline in HAI titers that occurred in participants 90-95 years of age (decrease of 1.65 units; p < 0.0001) (**Figure 5.I**). Here, the sex difference in pre-vaccination titers was only significant at the oldest end of the cohort (p = 0.003 at 95 years of age). Taken together, these data illustrate sex-specific effects of aging on pre-vaccination antibody titers to H3N2 and influenza B, but not H1N1.

### Controlling for sex leads to incorrect interpretations

The impact of sex as a biological variable is often ignored in biomedical research^42,43^. This is particularly true in the context of aging, where attention is typically directed to immunosenescence, failing to recognize that aging can have distinct effects on immune function in males and females^44-46^. In this analysis, *a priori* hypotheses regarding sex differences in the effect of age guided the inclusion of age by sex interaction terms in all models. To illustrate the consequences of ignoring sex as a biological variable, however, analyses were repeated controlling for sex rather than allowing age effects to vary by sex. For H3N2 and influenza B, where the effect of aging was found to be statistically significantly different in males as compared to females, the estimates derived by controlling for sex (black lines in **Figure 5E, F, H, I**) were not representative of either sex. In the linear models, for example, controlling for sex led to the incorrect inference that titers remain constant with age, while the interaction models demonstrate that this is false for both males and females.

In addition to conceptual reasons to interrogate sex differences, inclusion of interaction terms improved the goodness-of-fit of statistical models. Goodness-of-fit of eight models for each vaccine antigen were compared using Akaike’s Information Criterion (AIC) in **Table 4**, where lower values indicate better relative goodness-of-fit. For antibody titers to H3N2 and influenza B, despite the penalty for increasing model complexity, fit was improved by including an age-sex interaction term, and the best models overall were expanded models that allowed the effect of age to differ by sex. Thus, incorporating sex differences into vaccinology research can lead to more robust analysis.

**Table 4.**
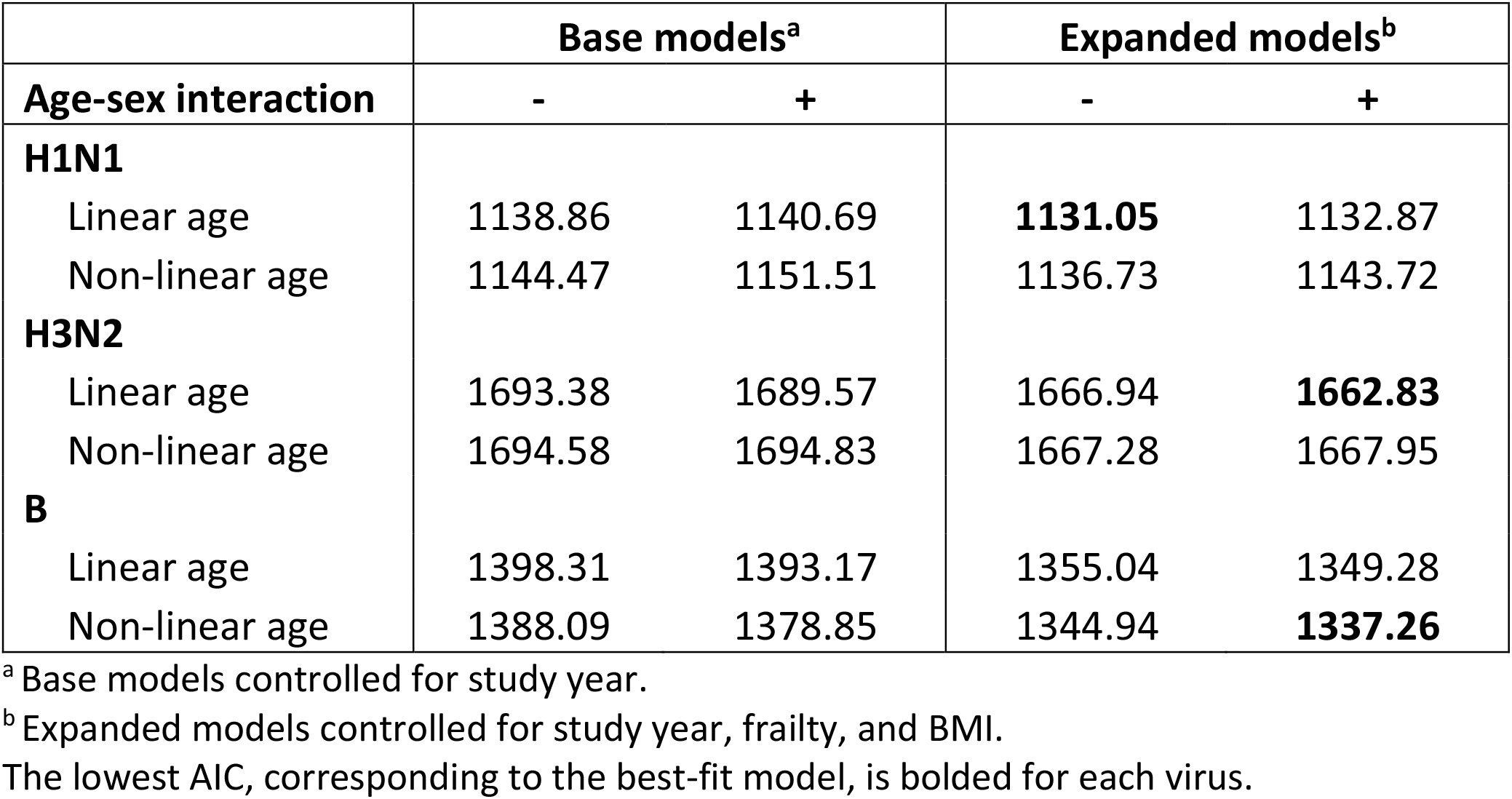
Goodness-of-fit comparison of pre-vaccination age models.

## Discussion

In this multi-season, longitudinal study of older adults over 75 years of age, we observed no statistically significant associations of age, sex, BMI, frailty, or the number of years of repeat annual vaccination with any post-vaccination HAI titer outcomes. Importantly, this suggests that repeat annual vaccination with HD-IIV3 does not seem to have significant positive or negative effects on strain-specific HAI antibody responses in the highly vaccinated elderly population but may mask the impact of other factors. Interestingly, there was some evidence to suggest that particularly for H3N2, the effect of repeat vaccination does vary by sex, thus supporting our hypothesis that variability in estimates of the effect of repeat vaccination in the literature is caused by failure to adequately account for the contribution of host factors.

Pre-vaccination titers strongly predicted all post-vaccination antibody titer outcomes, indicating that pre-existing humoral immunity, which may reflect the durability of humoral immunity against influenza from previous seasons, is an important outcome measure to consider in the highly vaccinated older adult population. Regarding age as a determinant of pre-existing humoral immunity, we found that HAI titers to all three vaccine antigens stayed constant in females with age but that HAI titers to H3N2 and influenza B decreased with age in males, leading to significant sex differences in the effect of age for these two viruses. It has previously been reported that at older ages, there is a male-bias in influenza B infection and hospitalization^47,48^. Our results thus provide a potential mechanism for this sex difference and highlight the need to develop better vaccines or vaccination strategies for older males. Finally, we demonstrated that failing to allow for sex-specific effects would have resulted in invalid conclusions for both males and females.

Because older adults are disproportionally burdened by severe disease and mortality from seasonal influenza, significant effort has been devoted to improving annual vaccination coverage for this vulnerable population. Older adults, particularly those who are over 75 years of age, can thus have decades of repeated annual influenza vaccination. Cumulatively, repeated annual vaccination can lead to high pre-vaccination titers. As expected, over 72% participants had a pre-vaccination HAI titer of 40 or higher to H1N1 and H3N2, and 98% participants had such “seroprotection” to influenza B. High pre-existing humoral immunity has important consequences in assessing humoral immune responses to influenza vaccines, as demonstrated by the strong positive relationship between pre- and post-vaccination titers. Conversely, ratio-based outcomes (i.e., fold rise and seroconversion) were negatively associated with pre-vaccination titers, since, by definition, large denominators (pre-vaccination titers) result in smaller ratios. We recently reported a similar effect in younger adult healthcare workers, where mandatory vaccination policies result in exceptionally high rates of immunization^49^. While some argue that high levels of pre-existing antibodies interfere with the generation of *de novo* immune responses^24,27^, others suggest that ratio-based measures simply do not appropriately control for imbalances in levels of pre-existing humoral immunity^50^. Following the latter interpretation, a “ceiling effect” is evident in our data, whereby the limit of detection of the assay prohibits observing seroconversion or a large fold-rise in individuals with high levels of pre-existing humoral immunity. Particularly important in older adults, where formation of *de novo* responses is impaired by immunosenescence^51^, the breadth of pre-existing humoral immunity and the positive predictive value for post-vaccination titers can thus be harnessed to elicit protection^26^. The clinical and scientific implications of this notion are far-reaching and long-term, as the Advisory Committee on Immunization Practices (ACIP) of the CDC has recommended annual influenza vaccination for anyone aged 6 months and older since 2010^52^.

For many vaccines, antibody titers wane over time^53-56^. Influenza vaccines are unique in this respect due to the recommendation for yearly immunization and exceptional antigenic diversity, which alter the dynamics of waning immunity. The constant pre-vaccination HAI titers with age in females seen in our study suggest that females benefit from a booster effect from each successive annual vaccination that appears to prevent antibody waning. This influenza-specific effect has been reported elsewhere, where samples collected from individuals over a 20-year period revealed longitudinal increases in neutralizing titers to influenza^57^. As an explanation for their findings, the authors suggest that even as viral antigens drift over time, many epitopes remain conserved. These conserved epitopes stimulate memory B cells that were generated in response to previous exposures, thus periodically boosting steady-state immunity^57^. However, our data suggest that this effect is absent in males for H3N2 and influenza B. The reasons for this sex difference are unknown, but may be attributable to the compounding effects of females developing stronger responses to influenza infection and vaccination throughout adulthood^58^, leading to a more robust repertoire of memory B cells that recognize conserved epitopes on drifted virus strains. It is speculated that in older adults, consistently inferior responses among males may manifest as a lack of memory B cells that can be boosted by drifted viruses to counteract waning of antibody over time, thus resulting in decreasing pre-vaccination titers with age.

Notably the sex difference was absent for H1N1. A possible explanation for this lies in the differing evolutionary rates of the three viral strains. H1N1 experiences slower evolution than H3N2^59^ and the influenza B/Victoria lineage^60^. In addition, the global co-circulation of B/Yamagata and B/Victoria lineages leads to increased exposure to divergent antigens^61^. Accordingly, over the six influenza seasons included in our study, vaccine antigens were significantly more variable for H3N2 and influenza B than for H1N1. While three H1N1 strains were included in seasonal vaccines, five H3N2 strains and four influenza B strains were included over the six study years. A decade-long review of the number of divergent vaccine strains that our study participants may have been exposed to indicates that since 2010, the number of H1N1 vaccine strains remained at a total of three, while seven H3N2 strains and five influenza B were used in trivalent vaccines over this period^62^. It is thus possible that repeated exposure to the same H1N1 antigen sufficiently boosted male steady-state immunity to mask sex differences in the immune response. Conversely, for H3N2 and influenza B, sequential exposure to drifted virus strains required robust and broad responses to allow for boosting of steady-state immunity, which may have only been present in females.

Another possible explanation is immunological imprinting in youth, as it has a lifelong impact on subsequent immune responses to influenza infection and vaccination^22,63^. Individuals in our cohort, born from 1916-1941, may have been exposed to H1N1 in their youth, while the 1918 pandemic virus continued to circulate, but were likely exposed to H3N2 and influenza B later in life^64^. Accordingly, older adults are more likely to have HAI activity against historical strains of H1N1 viruses than younger subjects, but also have more restricted reactivity to historical H3N2 viruses^65^. Older cohorts are also relatively protected against symptomatic H1N1 infection compared to younger cohorts, but this pattern is reversed for H3N2^66,67^. It is therefore possible that strong immune imprinting to H1N1 virus strains masked the sex differences otherwise observed for H3N2 and influenza B.

Our study had several strengths and limitations. First, this was an observational study that was not specifically designed to interrogate sex differences in the immune response to influenza vaccination. To overcome small yearly sample sizes, six influenza seasons were pooled together, and statistical methods were used to control for annual variation in vaccine viruses and repeated measurements on participants. The resulting multi-season nature of this work improves generalizability to future influenza seasons. Secondly, the humoral immune response to vaccination was solely based on strain-specific HAI titers, which are the standard in the field, but lack the functional quality of microneutralization assays^68^. Relying solely on serological samples also prohibited more in-depth mechanistic investigation at the cellular level. Third, the lack of racial diversity in our cohort must be noted, as it prohibited us from investigating race as a host factor of interest, which should be considered in future studies. Although the study lacked racial diversity, the cohort was diverse in terms of age at vaccination, allowing us to study effects in the ‘oldest’ old subset. Finally, a major strength of this study is the intersectional approach to this analysis, which allowed for interrogation of effects both between and within groups (i.e., between and among males and females), leading to a richer and more nuanced interpretation^69^.

In conclusion, we demonstrated that in highly vaccinated older adults, pre-vaccination HAI titers, rather than age, sex, BMI, frailty, or repeat vaccination, predict post-vaccination parameters of humoral immunity. Further, these pre-vaccination titers change with age in a sex-specific manner such that older males are particularly vulnerable to lower levels of pre-existing humoral immunity. Further research should focus on elucidating the mechanisms underlying this sex difference, as well as novel vaccination strategies to harness the breadth of pre-existing immunity in older adults and better protect this vulnerable population.

## Methods

### Study population and protocol

During the 2014-2015 to 2019-2020 influenza seasons, we enrolled community-dwelling older adults above 75 years of age who had not yet received a seasonal influenza vaccine. Individuals who had a history of allergic reaction to influenza vaccines or to eggs, were currently taking oral steroids or had worsening or new-onset of immune-modulating conditions were excluded. Study participants came to the Clinical Research Unit at Johns Hopkins Institute of Clinical and Translational Research on the Johns Hopkins Bayview Medical Center campus, or study visits were done at participants’ home as needed. A detailed medical history was obtained, vital signs were measured and frailty was assessed as per the Frailty Phenotype^37^. After a pre-vaccination blood draw, participants received HD-IIV3 (Fluzone®High-Dose Sanofi Pasteur, PA, USA). A second blood sample was collected 21 to 28 days after vaccine administration (**Figure 1**). To focus on the context of repeat annual vaccination, only individuals who participated for a minimum of 4 study years were included in this analysis.

### Ethics

Written, informed consent was obtained from all participants. The study protocol was approved by the Johns Hopkins School of Medicine Institutional Review Board. The study is registered on clinicaltrials.gov (NCT02200276).

### Hemagglutination inhibition assays

A validated HAI assay was used to quantify antibody titers against study vaccine antigens for the three vaccine strains (H1N1, H3N2 and B) in each year and were performed by Sanofi Pasteur as previously described^70^. Briefly, serum was incubated with type III neuraminidase to eliminate non-specific inhibitors and then with turkey red blood cells to adsorb non-specific agglutinins. Sera were then serially diluted in duplicate, beginning at a 1:10 dilution, and incubated with influenza virus (4 hemagglutination units/25µl). Turkey red blood cells were then added, and the titer defined as the highest dilution in which hemagglutination was inhibited.

### Definitions and categorization of predictor variables

Sex was used as a dichotomous variable based on self-report. Age was calculated based on the date of vaccination and used as a continuous variable. The frailty assessment was based on the presence or absence of five measurable characteristics: slowed motor performance (by walking speed), poor endurance and energy (by self-report of exhaustion), weakness (by grip strength), shrinking (by unintentional weight loss), and low physical activity (self-report)^29,37^. Participants with three or more out of these five characteristics were defined as frail, those with one or two as prefrail, and those with none as non-frail. BMI was calculated based on measured height and weight and used as a continuous variable. Study year refers to the influenza season (i.e., 2014-2015 to 2019-2020) and was used as categorical variable so as not to imply a linear relationship from year-to-year. Number of years of study participation was defined as the number of vaccines administered to an individual as part of the study and was used as a continuous variable ranging from 1 to 6.

### Outcome variables

Geometric mean titers were calculated both pre- and post-vaccination. For regression analysis, titers were transformed to a log_2_ scale to achieve an approximately normal distribution. The fold-rise in titer was calculated as post-vaccination titers divided by pre-vaccination titers, and log_10_ transformed to achieve a normal distribution. Seroconversion was defined as achieving a fold-rise ≥4 and used as a binary outcome. Seroprotection was defined as a titer ≥1:40 and used as a binary outcome.

### Statistical analysis

To account for repeated measures on participants, multi-level mixed effects models with random intercepts on the individual were used. Following standard risk factor analysis procedure, the contributions of host factors of interest were first assessed individually. Based on the *a priori* hypotheses of this analysis, fixed effects of the base models for post-vaccination outcomes adjusted for study year, pre-vaccination titers and included interaction terms to allow the effect of the host factor to differ for males and females. Fixed effects of the base models for pre-vaccination titers adjusted for study year and included interaction terms to allow effects to differ by sex. Where significant sex differences were found, further analysis controlled for additional covariates, and used cubic B-splines to investigate non-linear relationships^71^. The relative goodness-of-fit of various models were compared using Akaike’s Information Criterion. For graphs, predictions were capped at 95 years of age due to low sample size and large uncertainty in estimates above 95 years. Coefficients were considered statistically significant if 95% confidence intervals did not span the null value of zero (i.e., p<0.05). Analysis was performed in Stata 15 (StataCorp).

## Data Availability

The data that support the findings of this study are available from the corresponding author upon request.

## Acknowledgements

The authors thank the participants, as well as the clinical staff at the Healthy Aging Studies Unit and the Johns Hopkins University Institute for Clinical and Translational Research. The authors would also like to than Dr. Andrew Pekosz for helpful discussion and feedback. Sanofi Pasteur provided HD-IIV3 and performed HAI testing, but remained blinded to the results and had no further scientific input. This work was supported in part by National Institute of Health (NIH)/National Institute of Allergy and Infectious Diseases R01 AI108907 to S.X.L., NIH/National Institute on Aging Specialized Center of Research Excellence U54 AG062333 awarded to S.L.K., and funding from Irma and Paul Milstein Program for Senior Health, Milstein Medical Asian American Partnership (MMAAP) Foundation of USA to S.X.L. J.R.S was supported by a training award from the Fonds de recherche du Québec – Santé (File #287609). C.J.W, X.X.N, and S.F.L. were supported by Irma and Paul Milstein Program for Senior Health fellowship awards from MMAAP Foundation of USA (www.mmaapf.org).

## Author contributions

S.X.L. designed and oversaw the study. E.A., D.C.K. and E.S-M collected data and biological samples. H.F.L., Y.Y.C., Y.C., C.J.W., X.X.N., L.W., and S.F.L. processed biological samples. J.R.S., K.M., and R.S. entered and cleaned data. J.R.S. conceived of analytical approach and experimental questions and performed analyses under mentorship of S.L.Z, R.M., and S.L.K. K.M., H.K., and P.S. contributed to analyses. J.R.S. wrote the manuscript, with significant editorial contributions and discussion from S.L.K., R.M., S.L.Z., and S.X.L. All authors reviewed, edited, and approved the final draft of the manuscript. All authors are accountable for the accuracy and integrity of the work.

